# Insulin pricing and other major diabetes-related concerns in the USA: A study of 46,407 Tweets between 2017 and 2019

**DOI:** 10.1101/2020.01.04.20016527

**Authors:** Adrian Ahne, Francisco Orchard, Xavier Tannier, Camille Perchoux, Beverley Balkau, Sherry Pagoto, Jessica L Harding, Thomas Czernichow, Guy Fagherazzi

**Affiliations:** Inserm U1018, Center for Research in Epidemiology and Population Health (CESP), Paris Saclay University, Gustave Roussy Institute, 114 rue Edouard Vaillant, 94800 Villejuif, France; Epiconcept, 47 rue de Charenton, 75012 Paris, France; Inserm UMRS 1142, LIMICS, 15 rue de l’école de médecine, 75006 Paris, France; Sorbonne Université, 4 Place Jussieu, 75005 Paris, France; Luxembourg Institute of Socio-Economic Research (LISER), Esch/Alzette, 11 Porte des Sciences, 4366 Esch-sur-Alzette, Luxembourg; UConn Center for mHealth & Social Media, University of Connecticut, 2006 Hillside Road, Unit 1248, Storrs, CT 06269-1248, Connecticut, United States of America; Division of Transplantation, Department of Surgery, Emory University School of Medicine, Emory University Hospital, 101 Woodruff Circle NE, Suite 5127 Woodruff Memorial Research Bldg Atlanta, Georgia 30322 United States of America; Luxembourg Institute of Health; Department of Population Health, 1A-B, rue Thomas Edison, L-1445 Strassen, Luxembourg

**Keywords:** Emotion, Epidemiology, Psychological Stress, Methodology

## Abstract

**Introduction:** Little research has been done to systematically evaluate concerns of people living with diabetes through social media, which has been a powerful tool for social change and to better understand perceptions around health-related issues. This study aims to identify key diabetes-related concerns in the USA and primary emotions associated with those concerns using information shared on Twitter.

**Research Design and Methods:** A total of 11.7 million diabetes-related tweets in English were collected between April 2017 and July 2019. Machine learning methods were used to filter tweets with personal content, to geolocate (to the US) and to identify clusters of tweets with emotional elements. A sentiment analysis was then applied to each cluster.

**Results:** We identified 46,407 tweets with emotional elements in the USA from which 30 clusters were identified; five clusters (18% of tweets) were related to insulin pricing with both positive emotions (*joy, love)* referring to advocacy for affordable insulin and *sadness* emotions related to the frustration of insulin prices, five clusters (12% of tweets) to solidarity and support with a majority of *joy* and *love* emotions expressed. The most negative topics (10% of tweets) were related to diabetes distress (24% *sadness*, 27% *anger*, 21% *fear* elements), to diabetic and insulin shock (45% *anger*, 46% *fear*) and comorbidities (40% *sadness*).

**Conclusions:** Using social media data, we have been able to describe key diabetes-related concerns and their associated emotions. More specifically, we were able to highlight the real-world concerns of insulin pricing and its negative impact on mood. Using such data can be a useful addition to current measures that inform public decision making around topics of concern and burden among people with diabetes.

**Significance of Study:** *What is already known about this subject?:*

- It is very challenging to collect representative data at a population level to understand what are the key concerns of people with diabetes in real life.
- Social media platforms, such as Twitter, may serve as a relevant source of information to supplement traditional population health studies.
- There are worldwide inequalities in access to insulin.

*What are the new findings?:*

- With 18% of the tweets related to insulin pricing, this is a major concern in the diabetes community in the USA.
- People regularly express fear, anger and sadness about potential diabetes-related complications and comorbidities.
- However, there is a lot of support and solidarity among the diabetes online community, with numerous posts related to positive emotions

*How might these results change the focus of research or clinical practice?:*

- Our work presents a reproducible approach to easily capture information about key diabetes-related concerns, that is usually not available in typical clinical or epidemiological studies. This information can supplement data from clinical or epidemiological studies to inform public health strategies to deal with diabetes-related prevention, management and treatment

## INTRODUCTION

Stress, fears and negative emotions are considered to be the most important psychosocial health factor in the management of diabetes.^1^ Diabetes distress is associated with decisional conflict and therefore has an impact on day-to-day disease management and the long term risk of diabetes-related complications.^2^ Reducing diabetes-related distress may improve HbA1c and reduce the burden of disease among people with diabetes.^3,4^ However, there is still limited knowledge about the sources of stress, anxiety and concerns among people with diabetes and it is difficult to capture them with existing evaluation scales such as the Problem Areas in Diabetes Scale (PAID or PAID-T for adolescents) or the Diabetes Distress Scale (DDS).^5–9^

Social media data offer a unique opportunity to supplement current measures of diabetes-related distress and assess the sentiments of people with diabetes, given the very active online diabetes community in particular on Twitter.^10,11^ Twitter is a microblogging and social networking service with 321 million monthly active users.^12^

Using social media to explore psychological information related to diabetes and associations between socioeconomic factors and diabetes-related concerns based in real-life is currently unchartered territory. However, using this resource may provide important insights and allow future interventions regarding prevention, management and treatment of diabetes to be more appropriately tailored.

Using Twitter data, the aims of the current study are to identify diabetes-related concerns in the USA and to identify the primary emotions and sentiments associated with them.

## METHODS

### Data collection

A tweet extraction engine was developed in April 2017 and since then has collected more than 11.7 million (status July 2019) diabetes-related tweets in English, via Twitter’s Streaming Application Programming Interface (API), based on a list of diabetes-related keywords, such as *diabetes, hypoglycemia, hyperglycemia* and *insulin*, from all over the world (see Supplementary Materials S1 for the full list of keywords used). All data collected in this study were publicly posted on Twitter. Therefore, according to the privacy policy of Twitter, users agree to have this information available to the general public.^13^ In this study, we restricted the analysis to 46,407 US-based tweets, with personal and emotional content (see Figure 1 for an overview over the workflow).

**Figure 1:**
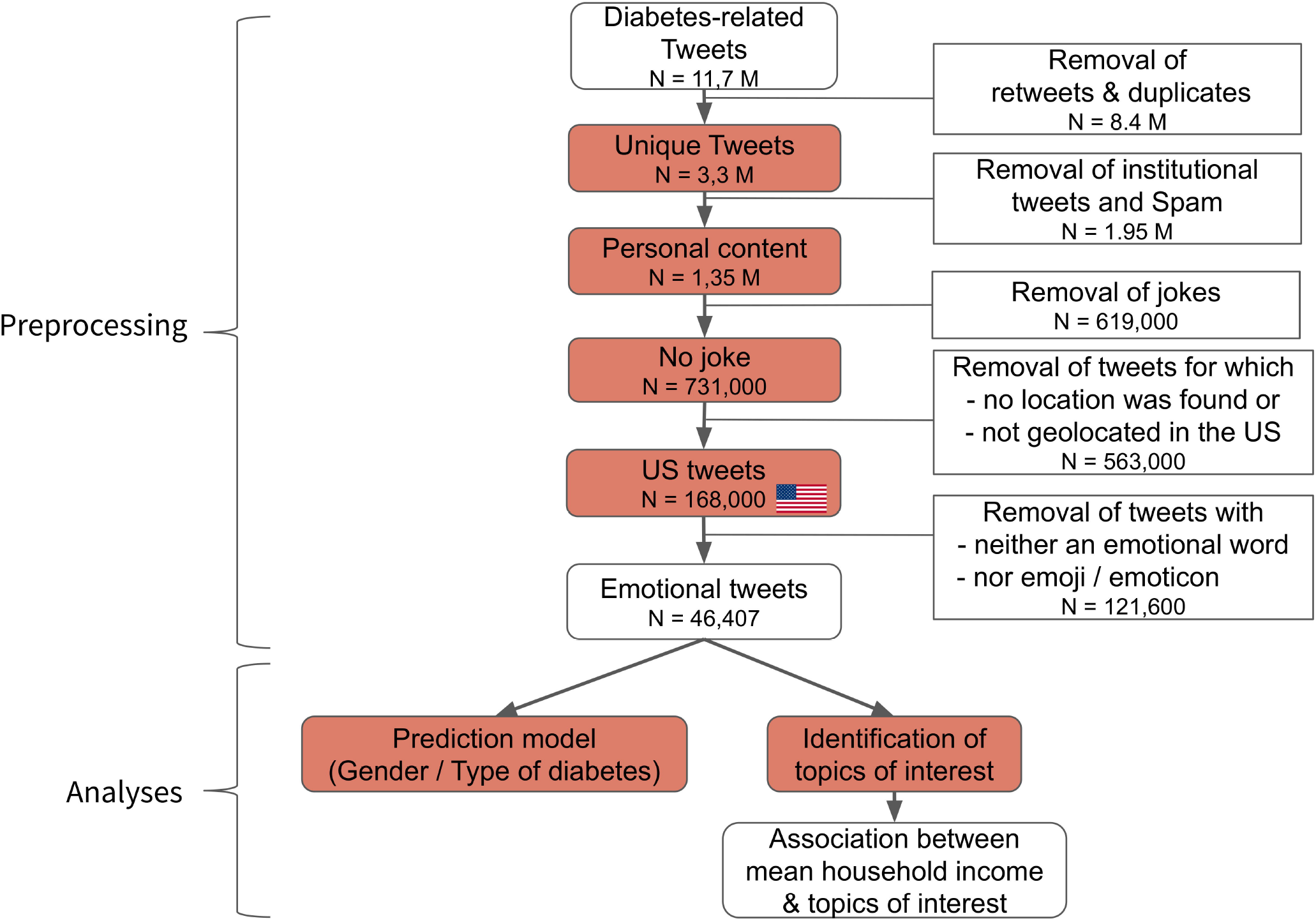
Workflow: from the data collection to the analysis.

For further details concerning data representation, data preprocessing, training of machine learning classifiers to filter tweets with personal content, from institutional tweets (organisations, advertisement, research news, etc.) related to diabetes; detect and exclude jokes; predict the gender and type of diabetes of users as well as the geolocation algorithm to determine tweets in the USA, please refer to Supplementary Material S2. For the present work, the statistical unit considered is a tweet and not a user profile. Thus a user may have several tweets that are included.

### Identifying emotions

To identify emotions among diabetes-related tweets, we combined detailed Parrot’s classification of emotions with a dictionary of emotional keywords present in the two most common tools to assess psychological health in people with diabetes, namely the Problem Areas in Diabetes questionnaire and the Diabetes Distress Scale.^14^ Parrot identified over 100 emotions and conceptualized them as a tree structured list (full list Supplementary Material S3) with the six primary emotions being *joy, love, surprise, sadness, anger* and *fear*. To capture as many tweets as possible containing emotions, we hypothesized that the synonyms of the words from Parrot’s classification should be included as well. Synonyms were identified using the WordNet database.^15^

Moreover, due to the nature of Twitter as a microblogging service (short messages), people use emoticons and emojis that express special meanings. Emoticons constitute a metacommunicative pictorial representation of a facial expression using punctuation marks and letters, such as ‘:-)’ or ‘:D’.^16^ Emojis are incorporated into sets of characters available in mobile phones, such as 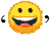 or 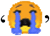. We used Wolny’s categorisation of emojis and emoticons into emotional categories similar to the six primary emotions defined by Parrot.^17^ This allowed us to determine a subset of *emotional* tweets containing either emotional words or emojis/emoticons.

### Sentiment Analysis

Sentiment analysis is commonly used to investigate the positive or negative opinion within a tweet. We used a widely accepted human-validated sentiment analysis tool Valence Aware Dictionary for Sentiment Reasoning (VADER).^18–21^ VADER was specifically developed for social media data and combines a lexicon and the processing of the sentence characteristics to determine sentence polarity. VADER computes sentiment and valence for each word level and provides positive, negative and neutral scores at the sentence level. We used the compound score as our main metric for the sentiment analysis (referred to as the *SA score* later in the text), which is unidimensional and a normalized measure of sentiment between −1 (most negative) and +1 (most positive).

### Topic extraction

All tweets are represented via their word vector representations, with the consequence that words similar in semantics are also similar in the word vector space (see Supplementary Material 2 for more details). We applied the unsupervised machine learning algorithm K-means, using cosine similarity as distance measure, to group the emotional tweets that are close to each other based on their word vector representations into topics/clusters.^22^ In order to define the right number of clusters *k*, the Silhouette score, measuring how on average each data point is closer to its cluster’s center than to any other cluster, was used as input parameter for the K-means algorithm.^23^ We obtained the highest score for *k=30* clusters. All tweets were then assigned to one of the 30 clusters/topics. Each topic/cluster was then given a label by two authors (AA, GF) according to the 10 most contributing tweets (those closest to the topic center) and the most frequent words (top words) in the cluster.

### Assessment of the mean income

We studied the associations between the topics of interest for people with or talking about diabetes and the mean income of the city, based on data from the 2017 American Community Survey.^24^ The tertiles for the mean household income were calculated: low income - [$24,609, $67,224], medium income - [$67,225, $86,758], high income - [$86,759, $394,259]. Each geolocated tweet was associated with the mean income for its corresponding city and then assigned to its respective tertile: low income, medium (med) income, high income. Associations between topics and mean income tertile groups were assessed with Chi-squared tests. For each topic the p-value was calculated using a Chi2 test between the binary variable if a tweet belongs to the corresponding topic and the tertile categories of the city-level mean income from the tweet’s origin.

Python (version > 3.6) and the packages scikit-learn (machine learning algorithms and data preprocessing methods) and gensim (text processing, word representation) were used to implement the methods described above.^25,26^ Algorithms related to the present study are open source under the following address: https://github.com/WDDS/Tweet-Diabetes-Classification.

## RESULTS

### Spatial distribution of diabetes-related tweets

This analysis was based on 167,743 geolocated, diabetes-related tweets from the USA, see Figure 2. The highest number of tweets were seen in California (N: 18,551) and Texas (N: 14,237), whereas Vermont (N: 197) and Wyoming (N: 131) had a low number of tweets. At the city level, New York City (N: 9,663), Los Angeles (N: 5,301) and Chicago (N: 4,884) had the highest number of tweets.

**Figure 2:**
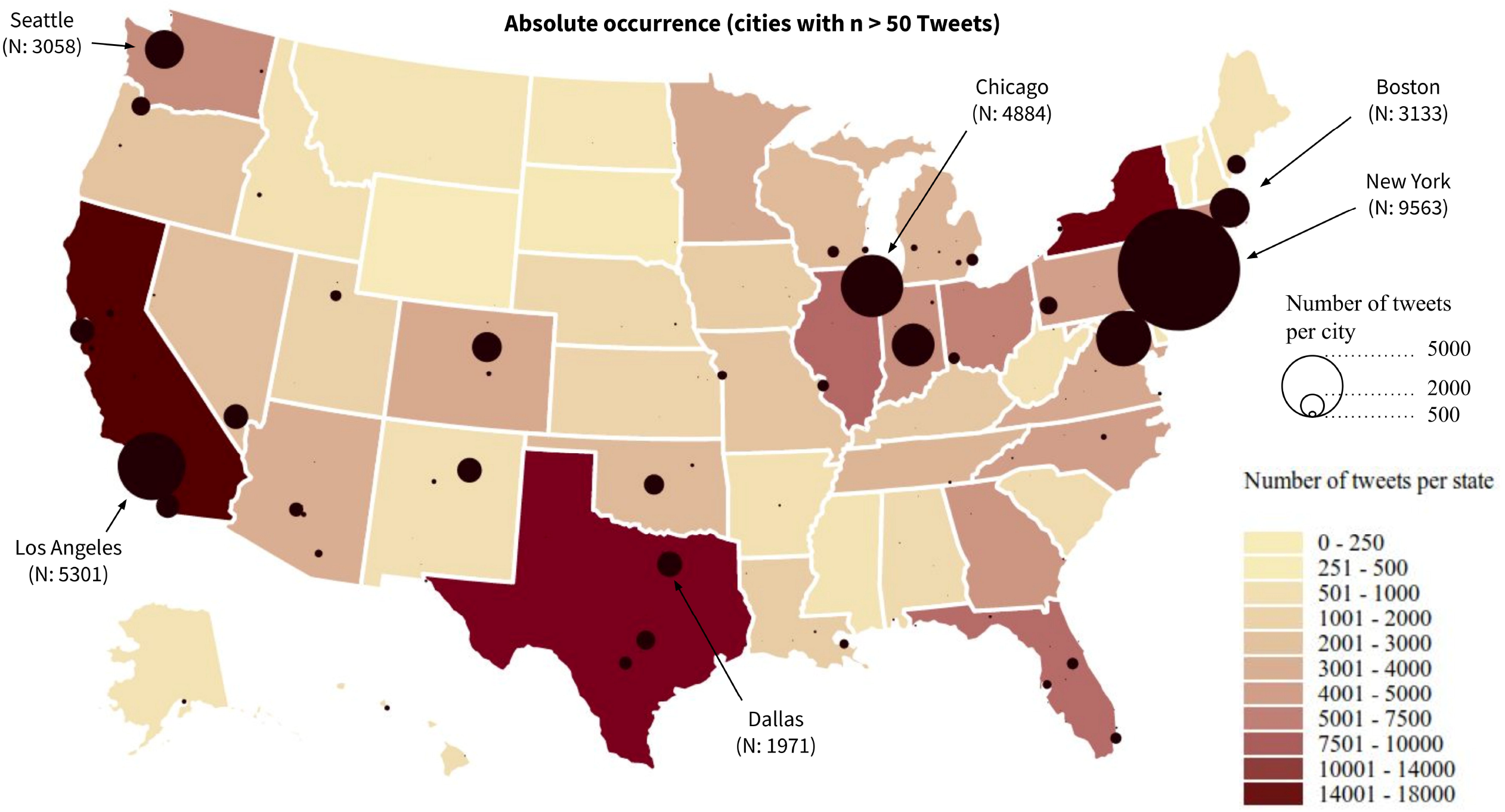
Spatial distribution of the diabetes-related tweets over the USA.

### Topics of interest

Among all US diabetes-related tweets, 46,407 were identified as *emotional* tweets (28% of tweets) of which 14,485 (31%) were written by men, 20,228 (44%) by women and 11,694 (25%) from unknown sex; 20,285 (44%) were predicted as from people with type 1 diabetes, 4,375 (9%) from type 2 diabetes and 21,747 (47%) diabetes type unknown. Table 1 shows the detailed description of the 30 topics of interest about diabetes.

**Table 1:**
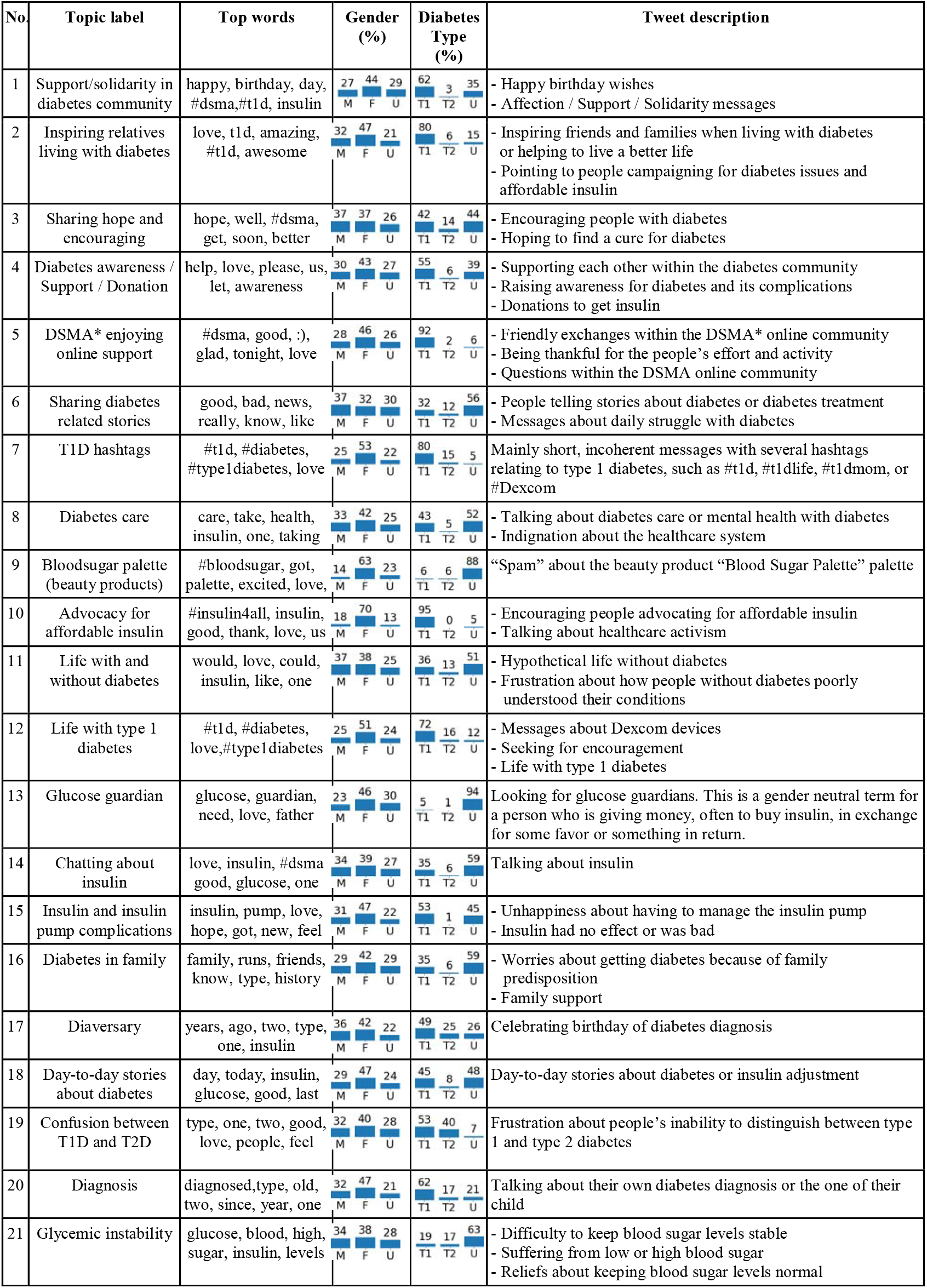
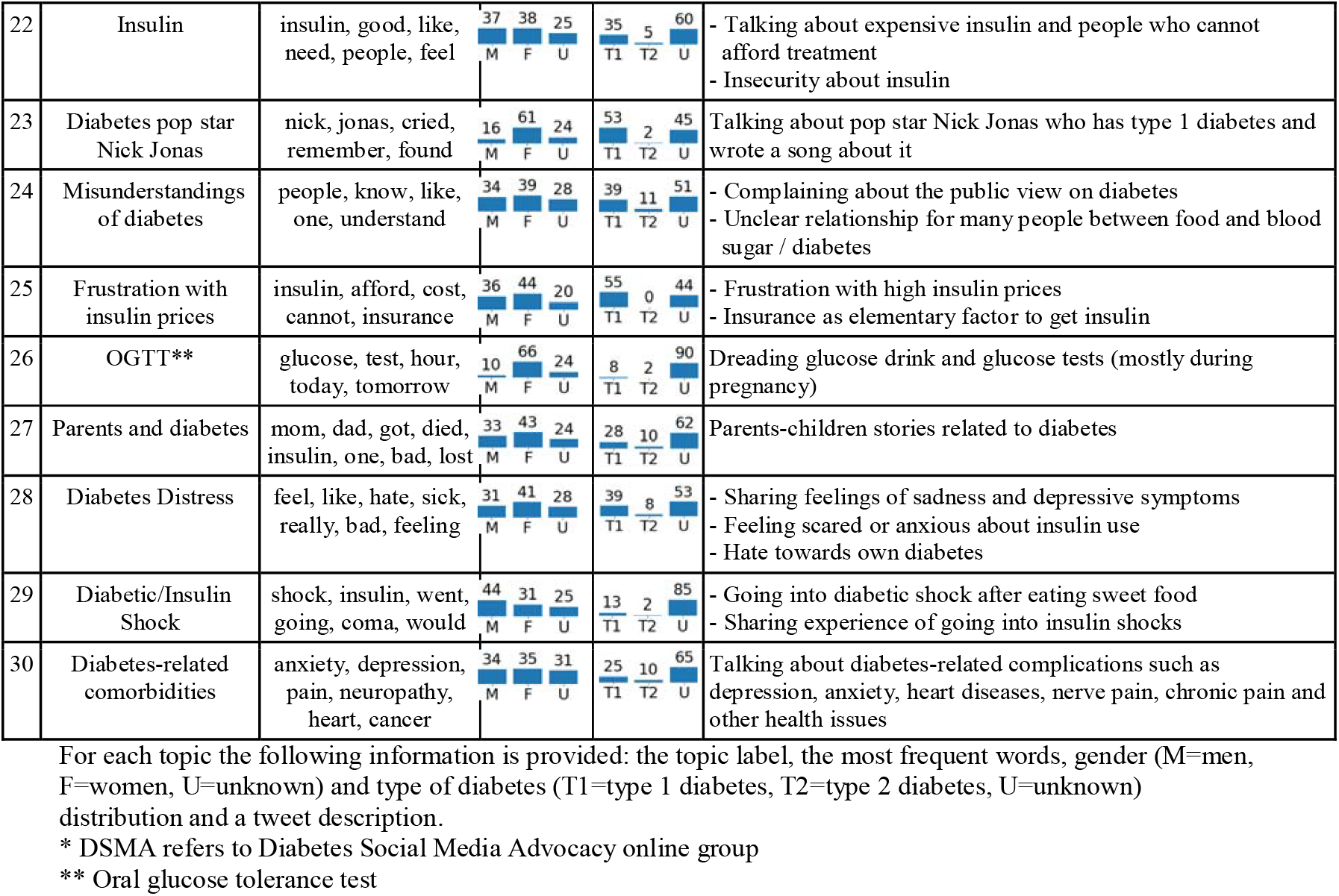
Overview of the 30 topics of interest for people with or talking about diabetes and their gender and type of diabetes distributions.

In most topics, an over-representation of women was observed, in particular in topics 10, 23 and 26 related to the importance of affordable insulin and the oral glucose tolerance test (OGTT). In contrast, men more frequently discussed topics around diabetes-related stories (topic 6) and ‘diabetic/insulin shock’ (topic 29). Discussions about ‘advocacy for affordable insulin’ (topic 10) and enjoying the exchange in the diabetes online community (topic 5) were almost exclusively dominated by people with type 1 diabetes. The only topics where people with type 2 diabetes had significantly more tweets than on average, is topic 19 related to the confusion between type 1 and type 2 diabetes or in topic 17 when they tweeted about the anniversary of their diagnosis. The Diabetes Social Media Advocacy (#DSMA) group on Twitter is a community of people exchanging about diabetes related topics, in which the price of insulin is a central concern.

### Primary emotions related to the topics of interest

Table 2 illustrates the sentiment and emotions distribution over the 30 topics.

**Table 2:**
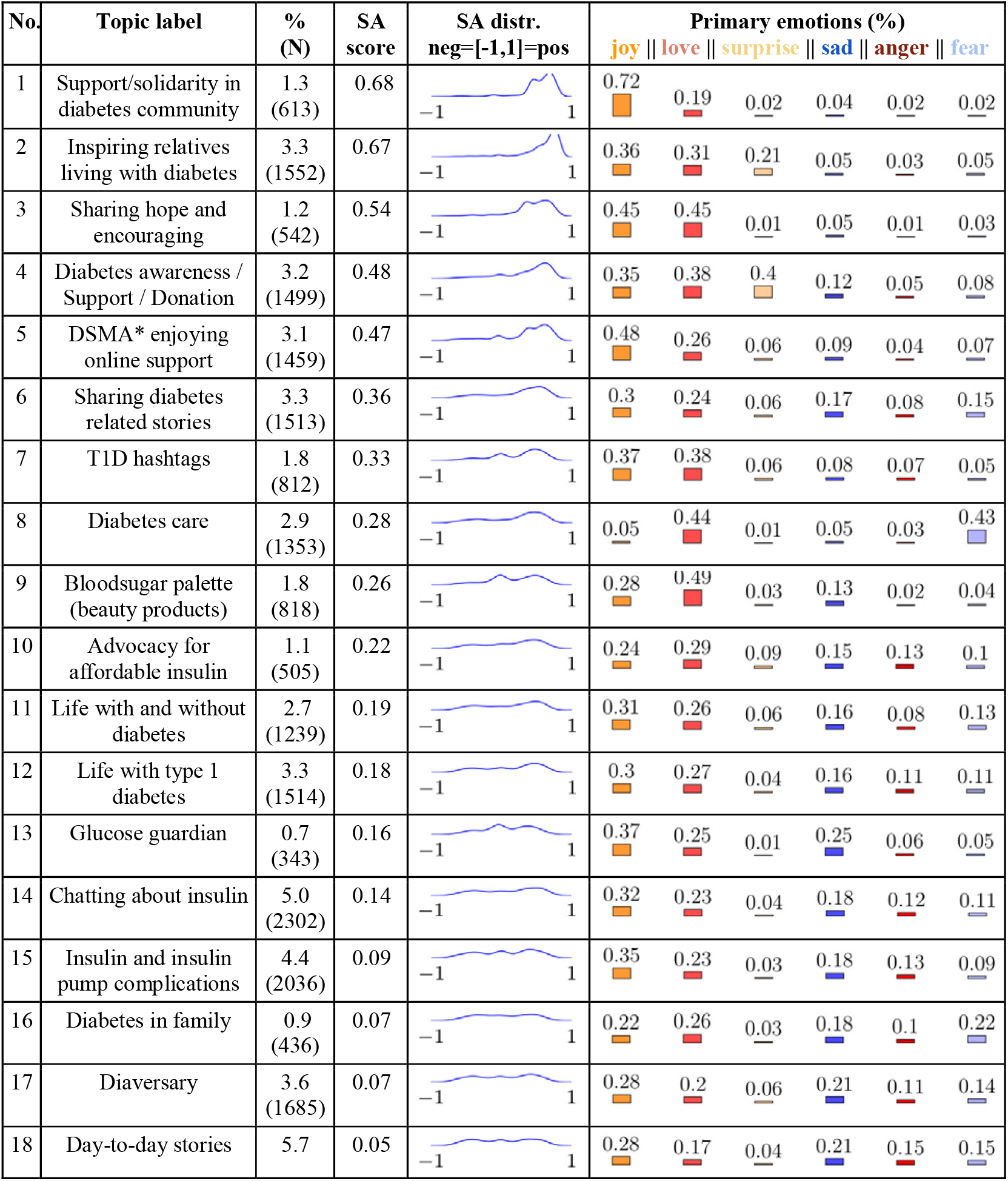
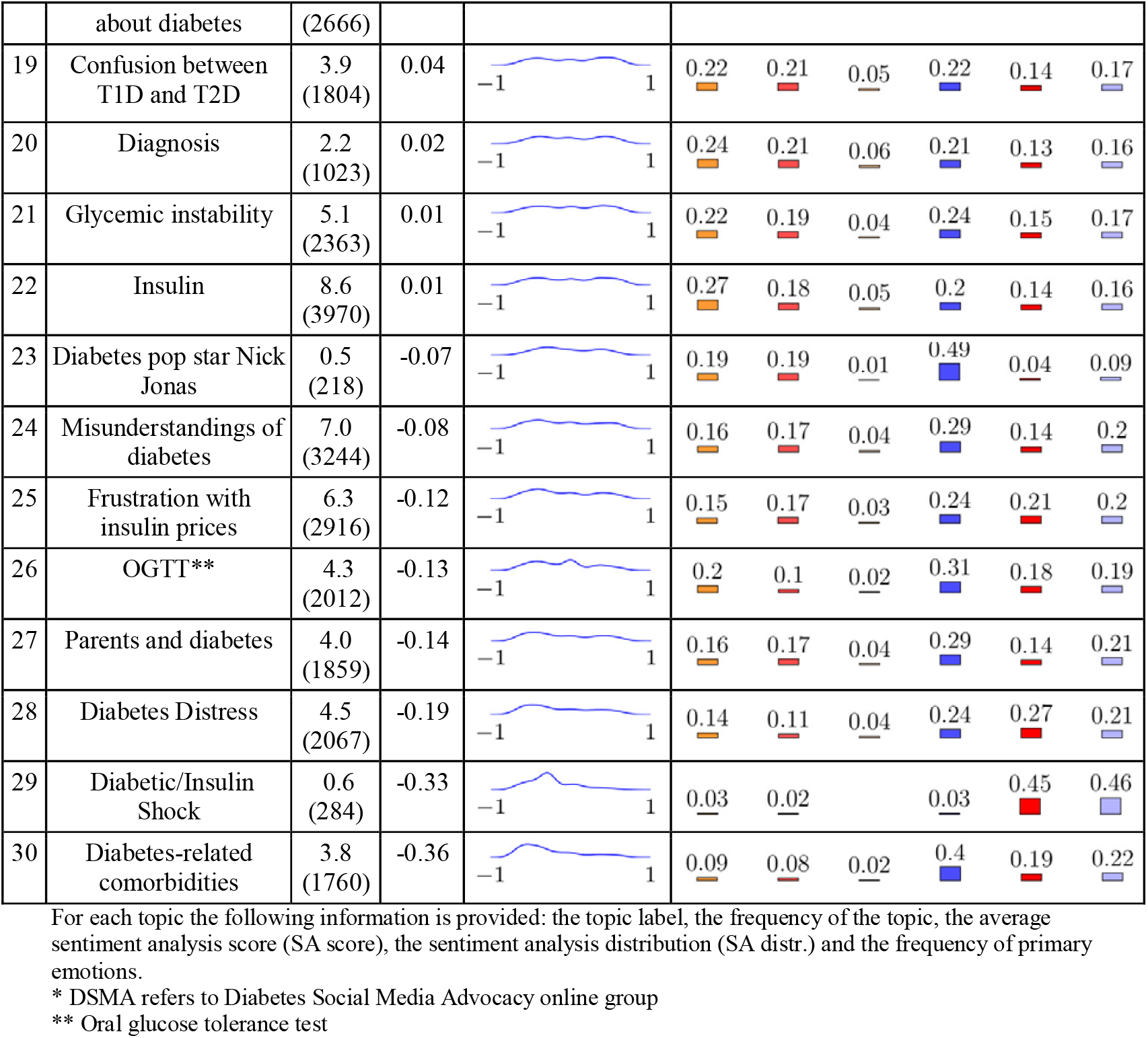
Sentiment and emotion distributions of the 30 topics of interest of people with or talking about diabetes.

| No. | Topic label | % (N) | SA score | SA distr.<br>neg=[-1,1]=pos | Primary emotions (%) |  |  |  |  |  |
| --- | --- | --- | --- | --- | --- | --- | --- | --- | --- | --- |
|  |  |  |  |  | joy | love | surprise | sad | anger | fear |
| 1 | Support/solidarity in diabetes community | 1.3 (613) | 0.68 |  | 0.72 | 0.19 | 0.02 | 0.04 | 0.02 | 0.02 |
| 2 | Inspiring relatives living with diabetes | 3.3 (1552) | 0.67 |  | 0.36 | 0.31 | 0.21 | 0.05 | 0.03 | 0.05 |
| 3 | Sharing hope and encouraging | 1.2 (542) | 0.54 |  | 0.45 | 0.45 | 0.01 | 0.05 | 0.01 | 0.03 |
| 4 | Diabetes awareness / Support / Donation | 3.2 (1499) | 0.48 |  | 0.35 | 0.38 | 0.4 | 0.12 | 0.05 | 0.08 |
| 5 | DSMA* enjoying online support | 3.1 (1459) | 0.47 |  | 0.48 | 0.26 | 0.06 | 0.09 | 0.04 | 0.07 |
| 6 | Sharing diabetes related stories | 3.3 (1513) | 0.36 |  | 0.3 | 0.24 | 0.06 | 0.17 | 0.08 | 0.15 |
| 7 | T1D hashtags | 1.8 (812) | 0.33 |  | 0.37 | 0.38 | 0.06 | 0.08 | 0.07 | 0.05 |
| 8 | Diabetes care | 2.9 (1353) | 0.28 |  | 0.05 | 0.44 | 0.01 | 0.05 | 0.03 | 0.43 |
| 9 | Bloodsugar palette (beauty products) | 1.8 (818) | 0.26 |  | 0.28 | 0.49 | 0.03 | 0.13 | 0.02 | 0.04 |
| 10 | Advocacy for affordable insulin | 1.1 (505) | 0.22 |  | 0.24 | 0.29 | 0.09 | 0.15 | 0.13 | 0.1 |
| 11 | Life with and without diabetes | 2.7 (1239) | 0.19 |  | 0.31 | 0.26 | 0.06 | 0.16 | 0.08 | 0.13 |
| 12 | Life with type 1 diabetes | 3.3 (1514) | 0.18 |  | 0.3 | 0.27 | 0.04 | 0.16 | 0.11 | 0.11 |
| 13 | Glucose guardian | 0.7 (343) | 0.16 |  | 0.37 | 0.25 | 0.01 | 0.25 | 0.06 | 0.05 |
| 14 | Chatting about insulin | 5.0 (2302) | 0.14 |  | 0.32 | 0.23 | 0.04 | 0.18 | 0.12 | 0.11 |
| 15 | Insulin and insulin pump complications | 4.4 (2036) | 0.09 |  | 0.35 | 0.23 | 0.03 | 0.18 | 0.13 | 0.09 |
| 16 | Diabetes in family | 0.9 (436) | 0.07 |  | 0.22 | 0.26 | 0.03 | 0.18 | 0.1 | 0.22 |
| 17 | Diaversary | 3.6 (1685) | 0.07 |  | 0.28 | 0.2 | 0.06 | 0.21 | 0.11 | 0.14 |
| 18 | Day-to-day stories | 5.7 | 0.05 |  | 0.28 | 0.17 | 0.04 | 0.21 | 0.15 | 0.15 |
|  | about diabetes | (2666) |  |  |  |  |  |  |  |  |
| 19 | Confusion between T1D and T2D | 3.9 (1804) | 0.04 |  | 0.22 | 0.21 | 0.05 | 0.22 | 0.14 | 0.17 |
| 20 | Diagnosis | 2.2 (1023) | 0.02 |  | 0.24 | 0.21 | 0.06 | 0.21 | 0.13 | 0.16 |
| 21 | Glycemic instability | 5.1 (2363) | 0.01 |  | 0.22 | 0.19 | 0.04 | 0.24 | 0.15 | 0.17 |
| 22 | Insulin | 8.6 (3970) | 0.01 |  | 0.27 | 0.18 | 0.05 | 0.2 | 0.14 | 0.16 |
| 23 | Diabetes pop star Nick Jonas | 0.5 (218) | -0.07 |  | 0.19 | 0.19 | 0.01 | 0.49 | 0.04 | 0.09 |
| 24 | Misunderstandings of diabetes | 7.0 (3244) | -0.08 |  | 0.16 | 0.17 | 0.04 | 0.29 | 0.14 | 0.2 |
| 25 | Frustration with insulin prices | 6.3 (2916) | -0.12 |  | 0.15 | 0.17 | 0.03 | 0.24 | 0.21 | 0.2 |
| 26 | OGTT** | 4.3 (2012) | -0.13 |  | 0.2 | 0.1 | 0.02 | 0.31 | 0.18 | 0.19 |
| 27 | Parents and diabetes | 4.0 (1859) | -0.14 |  | 0.16 | 0.17 | 0.04 | 0.29 | 0.14 | 0.21 |
| 28 | Diabetes Distress | 4.5 (2067) | -0.19 |  | 0.14 | 0.11 | 0.04 | 0.24 | 0.27 | 0.21 |
| 29 | Diabetic/Insulin Shock | 0.6 (284) | -0.33 |  | 0.03 | 0.02 |  | 0.03 | 0.45 | 0.46 |
| 30 | Diabetes-related comorbidities | 3.8 (1760) | -0.36 |  | 0.09 | 0.08 | 0.02 | 0.4 | 0.19 | 0.22 |

For each topic the following information is provided: the topic label, the frequency of the topic, the average sentiment analysis score (SA score), the sentiment analysis distribution (SA distr.) and the frequency of primary emotions.
\* DSMA refers to Diabetes Social Media Advocacy online group
\*\* Oral glucose tolerance test

In Supplementary Material S4, the most common emotional words and emojis/emoticons have been listed for each topic. Overall, we found that the most positive topics were related to support and solidarity within the diabetes online community (topic 1) with 72% of emotions related to *joy* and about inspiring relatives living with diabetes (topic 2) with *joy* (36%) and *love* (31%) elements. In contrast, the most negative topics were related to ‘diabetes-related comorbidities’ (topic 30) of which 40% of the emotions were related to *sadness*, 19% to *anger* and 22 % to *fear*; ‘diabetic/insulin shock’ (topic 29) with an over-representation of *anger* (45% of all emotions) and *fear* (46%); and “diabetes distress” (topic 28) with *sadness* (24%), *anger* (27%) and *fear* (21%) emotions expressed. Topics 2 (“Inspiring relatives living with diabetes”) and 4 (“Diabetes awareness / Support / Donation”) showed a higher percentage of *surprise* emotions. The topics in which people with or talking about diabetes exchange, support each other and show solidarity (topics 1, 2, 3, 4, 5) contained *joy* and *love* emotions most frequently. Topic 9 was related to beauty products with diabetes-related names and was considered irrelevant for our analyses and disregarded.

### Insulin pricing

Insulin pricing is a major topic of interest mentioned in 5 of the 30 topics (18% of tweets). A key topic associated with insulin pricing was related to the frustration of not being able to afford insulin (topic 25), in which many tweets expressed sadness (24%), anger (21%) and fear (20%). Topic 13 regrouped tweets in which people are looking for “glucose guardians”, a gender neutral term for a person who is giving money, often to buy insulin, in exchange for some favor or something in return. Topic 10 described the tweets of people who are fighting for affordable insulin, which are represented by the hashtag ‘#insulin4all’. The positive emotions in this group showed mutual support.

### Associations between topics of interest and mean income

We observed that topics such as 10 ‘advocacy for affordable insulin’, 22: ‘insulin’ and 25: ‘frustration with insulin prices’ were positively associated with mean household city income such that cities with higher incomes were more likely to post tweets about these topics (P<0.001 for all) (see Supplementary Material S5). Similarly, we found that topics 24: ‘misunderstandings of diabetes’ (P<0.001), 19: ‘confusion between type 1 and type 2 diabetes’ (P<0.05), 12: ‘life with type 1 diabetes’ (P<0.01) and 21: ‘Glycemic instability’ (P<0.05) were positively associated with mean city income. Likewise, positive associations were found between topics 24 and 19 which are related to language use and the way people talk about type 1 and type 2 diabetes, with cities with higher incomes being more likely to talk about these subjects.

In contrast, we observed that topics 5: ‘diabetes social media advocacy’ (#DSMA) group enjoying online support (P<0.001), 18: ‘day-to-day stories about diabetes’ (P<0.001) and 26: ‘oral glucose tolerance test (OGTT)’ (P<0.001) were negatively associated with mean city income, such that cities with lower income were more likely to post these tweets. Furthermore, cities with medium mean income tweet about topic 15: ‘Insulin and insulin pump complications’, were more frequently (P<0.001).

## DISCUSSION

Our findings suggest that Twitter is a useful tool to capture key diabetes-related topics and the emotions associated with those topics. Our key findings suggest that there is a lot of support and solidarity among the diabetes online community with numerous tweets related to *joy* and *love* emotions observed. In contrast, Twitter users expressed fear, anger and sadness related to insulin pricing and diabetes-related complications and comorbidities, as well as considerable frustration about the inability of people to distinguish between type 1 and type 2 diabetes.

To our knowledge, this is the first study using social media data to capture information regarding key diabetes-related concerns. We are, therefore, unable to compare and contrast the results of this study with others. Nonetheless, the importance of understanding emotions and self-control (regulation of thoughts, emotions, and behavior) for health outcomes in people with diabetes has been previously documented in several other studies.^27^ Hagger et al. reviewed diabetes distress among adolescents with type 1 diabetes and found a substantial proportion experienced elevated diabetes distress and that it is often associated with suboptimal glycemic control.^28^ Richman et al. also suggested that positive emotions such as hope and curiosity may play a protective role in the development of disease.^29^ Ogbera et al. showed that higher levels of emotional distress are associated with poor self-care in type 2 diabetes.^30^ Another study conducted by Iturralde et al. showed that anxiety is highly comorbid with depression among individuals with type 2 diabetes.^4^ Our study aligns with these results showing that emotions and diabetes distress topics are frequent and concern people with diabetes or people talking about diabetes on social media.

Similar to Nguyen et al., who showed that individuals living in zip codes with high percentages of happy and physically-active tweets had lower obesity prevalence based on geolocated Twitter data, we have shown gradients between topics related to diabetes on Twitter and the household income level of their city.^31^

### Insulin pricing

We found that insulin pricing was a major concern among tweets shared in the US (18% of all tweets were related to insulin pricing) and is associated with both positive (*joy, love*) and negative (*sadness, anger, fear)* emotions. People frequently shared their frustration with insulin prices, access to insulin, and identifying sources of insulin including “glucose guardians” or donations, which represent major obstacles for people with diabetes.^32,33^ Positive emotions are present when it comes to solidarity in the fight for affordable insulin in the community. We observed associations of topics addressing insulin pricing to be more frequent in cities with high mean incomes. This does not necessarily indicate that people living in cities with a high mean household income feel more concerned about insulin prices, but rather they probably have a greater ability to tweet around this issue. A large number of tweets geolocated in cities with a high mean household income included the hashtag ‘#insulin4all’, a campaign that unites the diabetes community around the access to treatment for everyone.^34^

It is known that there are key challenges for a global and fair access to insulin.^35^ With respect to insulin pricing, we are the first to exhibit and quantify, on a large sample of people with or talking about diabetes, the extent of the crisis in the USA based on social media data. In addition, we have also been able to highlight the different emotions and fears associated with the crisis around insulin pricing.

### Strengths and Limitations

This study has numerous strengths. First, a major advantage of using social media data is that information is expressed spontaneously, on a large scale, and in real-time, in what can be considered as an open digital space with flat role hierarchy for information sharing and development of online communities. This potentially minimizes biases such as responder bias that you would observe in traditional and observational studies. We evaluated tweets related to diabetes from a large number of people with a large variability in their profiles. The methodologies developed in this study present an innovative way to concentrate on relevant (personal, emotional) geolocated tweets (US), to identify topics of interest and emotions shared within topics. This approach is able to capture trends in the online diabetes community as well as socioeconomic factors that can be associated with social media data at the ecological level. This new way of capturing data supplements the detection of topics which are less medically oriented.

There are, however, several limitations to consider. First, diabetes-related concerns expressed on Twitter may not be representative of all people with diabetes. However, it has been previously suggested that it can be partially offset by the large variability in the social media profiles, a key strength in digital epidemiology.^36^ While we did observe large variability in our Twitter profiles, we found an over-representation of people with type 1 diabetes and women in our study when compared to known diabetes epidemiology literature.^36^ The greater representation of type 1 diabetes may be explained by the younger demographics of Twitter users.^37^ Alternatively, type 1 diabetes may have more involved care, more devices, more challenging medication and more frustrations to report on Twitter as compared with type 2 diabetes. Regardless, our results should be interpreted in the context of the Twitter population only. Second, the precision of our filter classifiers (personal content, jokes), gender and type of diabetes classifiers, is not perfectly accurate, which means that we cannot guarantee that 100% of tweets are posted by actual people with diabetes and it was often impossible to define the sex or type of diabetes. Third, we were unable to account for several clinical and environmental factors that may help to tease out these associations. The label provided by the researchers for each topic is not exclusive. By refining the tweets in each topic, more subtopics could be defined. This could be a future direction to investigate. Fourth, the geolocation of tweets was partially based on locations the users provided, which might not be their true location. Fifth, emotion detection is still a challenge in the machine learning field due to the occurence of sarcasm and irony. It is one of the open research questions. Last, causal inference between the mean household income per city and the topics of interest of people residing in the corresponding city cannot be made as it is subject to ecological fallacy.

A perspective of our work is to extend our analyses to include more countries and languages.

## Conclusion

In this study we investigated diabetes-related topics and their associated emotions. We showed that insulin pricing is a central concern and comes with feelings of sadness, anger and fear. We have shown that using social media posts to capture emotions and concerns of individuals in real life is feasible, and is an efficient way of augmenting psychosocial, behavioral and epidemiological research. Our work should encourage future studies to consider social media data as supplementary information.

Social media provides a useful observatory for diabetes issues, as it is a direct source to capture information about people with diabetes feelings, emotions, beliefs and fears related to diabetes, diabetes treatment and complications among the large and active diabetes online community. The use of Twitter analysis on diabetes could inform the public debate about diabetes issues and help to contribute directly to public and clinical decision making. Social media data will help develop policies and interventions that consider key concerns among people with diabetes to ultimately improve health outcomes.

## Data Availability

All code is publicly available in this github repository: https://github.com/WDDS/Tweet-Diabetes-Classification
Data is available upon reasonable request

## Acknowledgment

Guy Fagherazzi takes full responsibility for the work as a whole, and for the decision to submit and publish the manuscript.

The authors’ contributions were as follows: G.F. designed the research; A.A. and G.F. conducted the research; F.O. and A.A collected the data; A.A., F.O. and G.F. analyzed data; A.A., G.F., F.O., T.C, X.T interpreted the data; A.A. and G.F. drafted the article; G.F., F.O., T.C., X.T., B.B., J.L.H., C.P. and S.P. revised the manuscript critically; GF had primary responsibility for the final content of the manuscript. All authors read and approved the final manuscript.

## License

The Submitting Author accepts and understands that any supply made under these terms is made by BMJ to the Submitting Author unless you are acting as an employee on behalf of your employer or a postgraduate student of an affiliated institution which is paying any applicable article publishing charge (“APC”) for Open Access articles. Where the Submitting Author wishes to make the Work available on an Open Access basis (and intends to pay the relevant APC), the terms of reuse of such Open Access shall be governed by a Creative Commons licence – details of these licences and which <u>Creative Commons</u> licence will apply to this Work are set out in our licence referred to above.

